# Comparison of fluid resuscitation strategies in hemorrhagic shock: Crystalloid vs Blood products

**DOI:** 10.1101/2025.03.17.25324130

**Authors:** Ghina Abdullah, Muhammad Ahmed Nadeem, Kashan Sikandar, Muhammad Sheharyar, Hafiza Khadija Shahid, Rahmah Ashar Sakrani, Shahzad Mahmood

**Author notes:** **Corresponding Author** Dr. Shahzad Mahmood.

## Abstract

**Background:** Hemorrhagic shock remains a leading cause of morbidity and mortality in trauma patients. The choice of resuscitation fluids plays a critical role in patient outcomes, with crystalloid solutions and blood products being the most commonly used strategies. This study aims to compare the efficacy of crystalloid versus blood product resuscitation in patients with hemorrhagic shock.

**Methods:** A sample of 30 dogs with hemorrhagic shock was randomized into two groups: the control group (crystalloid resuscitation) and the non-control group (blood product resuscitation). Key outcomes measured included hemodynamic stability, organ function (SOFA score), complications (fluid overload, coagulopathy), length of hospital stay, and recovery time. Data were analyzed to assess the effectiveness of each resuscitation strategy.

**Results:** The blood product group showed superior outcomes in terms of survival, hemodynamic stability, and organ function. The mean blood pressure was significantly higher post-resuscitation in the blood product group compared to the crystalloid group. Additionally, organ function improved faster in the blood product group, with lower SOFA scores at 48 hours. Complications such as fluid overload and dilutional coagulopathy were more common in the crystalloid group. The length of hospital stay and recovery time were also shorter in the blood product group.

**Conclusion:** Blood product resuscitation is more effective than crystalloid resuscitation in improving hemodynamic stability, organ function, and minimizing complications in patients with hemorrhagic shock. These findings support the use of blood products as the primary resuscitation strategy in severe hemorrhagic shock, although further research with larger sample sizes is necessary to confirm these results.

## Introduction

### Background and Rationale

Hemorrhagic shock is a critical condition that occurs when there is significant blood loss, leading to inadequate tissue perfusion and oxygenation, which can result in multi-organ failure and death if not promptly treated. It is one of the leading causes of morbidity and mortality in trauma patients, accounting for approximately 30% of trauma-related deaths worldwide.(2) The goal of fluid resuscitation in hemorrhagic shock is to restore circulating blood volume, improve oxygen delivery to tissues, and correct metabolic disturbances while minimizing the risk of complications such as coagulopathy, fluid overload, and organ damage.

Two primary strategies for fluid resuscitation in hemorrhagic shock are the use of **crystalloids** (e.g., Normal Saline, Lactated Ringer’s Solution) and **blood products** (e.g., packed red blood cells, fresh frozen plasma, and platelets).(3) Crystalloids, which are intravenous solutions containing water, electrolytes, and sometimes glucose, are widely used due to their availability, low cost, and ease of administration.(4) They are generally effective in restoring intravascular volume in the early stages of hemorrhagic shock. However, the use of large volumes of crystalloids can lead to dilutional coagulopathy, tissue edema, and inadequate oxygen delivery, particularly in cases of severe blood loss. (5)

Blood products, on the other hand, provide more comprehensive resuscitation by replacing lost components of blood, including red blood cells, clotting factors, and platelets. This makes blood products particularly valuable in managing severe hemorrhagic shock, where the loss of both volume and oxygen-carrying capacity is critical.(6) In contrast to crystalloids, blood products have been shown to improve tissue oxygenation, prevent coagulopathy, and reduce the risk of organ failure, but they come with their own set of complications, such as transfusion reactions, blood-borne infections, and a higher risk of fluid overload.

Despite the clinical advantages of blood products in severe hemorrhagic shock, there is still considerable debate surrounding the most effective resuscitation strategy, especially in patients with mild to moderate blood loss. Crystalloids are often used as the first line of treatment due to their wide availability and lower cost, but they may not provide the same benefits as blood products in terms of oxygen delivery and coagulation support. The optimal approach for fluid resuscitation in hemorrhagic shock remains a subject of ongoing research, particularly with respect to the balance between crystalloids and blood products and their respective roles in different stages of shock.

In light of these considerations, the present study seeks to compare the effectiveness of crystalloid solutions and blood products in the management of hemorrhagic shock. By evaluating clinical outcomes such as survival rate, organ function, and complications, this research aims to provide valuable insights into the relative benefits and risks of these two fluid resuscitation strategies. This study is particularly important in the context of trauma care, where timely and effective resuscitation can mean the difference between life and death. The study conducted on dogs will provide directions for human intervention.

## Research Objective

The primary objective of this research is to **compare the effectiveness of crystalloid solutions and blood products in the resuscitation of dogs with hemorrhagic shock and its** implication **to treat humans**. Specifically, the study aims to evaluate the following key outcomes:

1. **Hemodynamic Stability**: To evaluate and compare the improvements in vital signs (e.g., blood pressure, heart rate) in both treatment groups.
2. **Organ Function**: To assess the preservation of organ function, particularly renal, hepatic, and pulmonary function, through the use of the **Sequential Organ Failure Assessment (SOFA)** score.
3. **Complications**: To compare the incidence of complications, including fluid overload, dilutional coagulopathy, transfusion reactions, and organ dysfunction, between the two groups.
4. **Length of Hospital Stay and Recovery Time**: To evaluate the differences in the duration of hospital stay and recovery time in both treatment groups.

### Research Questions

1. How do crystalloid and blood product resuscitation strategies compare in terms of hemodynamic stability (e.g., blood pressure, heart rate) during and after resuscitation?
2. What is the impact of crystalloid vs blood product resuscitation on the preservation of organ function in dogs with hemorrhagic shock?
  ∘ Specifically, how do these resuscitation strategies affect renal, hepatic, and pulmonary function, as measured by the SOFA score?
3. What are the incidence and types of complications associated with crystalloid resuscitation compared to blood product resuscitation in hemorrhagic shock patients?
  ∘ This may include complications like fluid overload, coagulopathy, transfusion reactions, and organ dysfunction.
4. Does the use of blood products or crystalloids affect the length of hospital stay and recovery time for patients with hemorrhagic shock?

### Literature Review

#### Hemorrhagic Shock and Fluid Resuscitation

Hemorrhagic shock is a life-threatening condition that occurs when there is a significant loss of blood volume,(7) leading to inadequate tissue perfusion and oxygenation. It is a leading cause of mortality in trauma patients, particularly in cases of severe bleeding. The primary goal of fluid resuscitation in hemorrhagic shock is to restore the intravascular volume, maintain organ perfusion, (8) and correct the physiological disturbances caused by blood loss. In clinical practice, resuscitation strategies often include the use of **crystalloids** and **blood products**, both of which have been widely studied and debated for their effectiveness in treating hemorrhagic shock.

#### Crystalloid Resuscitation

Crystalloid solutions, such as **normal saline (NS), Lactated Ringer’s Solution (LR)**, and **Plasmalyte**, are commonly used as first-line resuscitation fluids (9) due to their widespread availability, cost-effectiveness, and ease of administration. They contain water and electrolytes, which help to restore circulating volume and improve hemodynamic stability in patients with hemorrhagic shock. However, while crystalloids are effective at expanding the intravascular volume, their role in improving tissue oxygenation and preventing organ dysfunction remains contentious.

A key issue with crystalloids is their tendency to cause **fluid shifts** and **dilutional coagulopathy**. Studies have shown that large volumes of crystalloids can lead to **third-space fluid accumulation**,(10) which is the retention of fluid in the interstitial or extravascular space, increasing the risk of pulmonary edema and tissue swelling. The **CRISTAL study** (2015) highlighted that although crystalloids were associated with similar survival outcomes as colloids, patients who received excessive crystalloid fluids often experienced complications such as tissue edema and delayed recovery of organ function.(11)

Additionally, while crystalloids are beneficial in expanding volume rapidly, they lack **oxygen-carrying capacity**(12) and do not address the root causes of hemorrhagic shock, such as **oxygen delivery and clotting**. The **fluid responsiveness** of crystalloids is limited, as they do not provide essential components for tissue oxygenation and clotting. Thus, while crystalloids remain an essential component of initial resuscitation, their use in patients with severe hemorrhagic shock is often questioned.

#### Blood Product Resuscitation

In contrast to crystalloids, blood products are resuscitation fluids that offer a more comprehensive solution to hemorrhagic shock. Blood products, such as **packed red blood cells (PRBCs), fresh frozen plasma (FFP)**, and **platelets**, provide oxygen-carrying capacity and coagulation factors that are lost during blood loss. They are particularly useful in managing severe hemorrhagic shock, where blood loss exceeds the capacity of crystalloids to restore volume and oxygenation effectively.

The use of blood products in hemorrhagic shock has been associated with **improved tissue oxygenation, reduced mortality**, and **better preservation of organ function**. A landmark study by **Hess et al. (2008)** found that patients who received early transfusions of blood products had improved survival rates and fewer complications, including less organ dysfunction and better hemodynamic stability. (13) The **PROPPR trial (2015)** demonstrated that in trauma patients with severe hemorrhagic shock, the early use of a balanced transfusion strategy with a 1:1:1 ratio of PRBCs, FFP, and platelets was associated with a significant reduction in mortality compared to a more traditional strategy using PRBCs alone. (14)

Blood products not only help with **oxygen delivery** but also address coagulopathy, which is a common complication in hemorrhagic shock. **Coagulopathy**, or the inability to form blood clots, is frequently observed in trauma patients due to the loss of clotting factors and platelets during bleeding. Blood products replenish these essential components, restoring the ability to form stable clots and control bleeding. This makes blood product transfusion particularly advantageous in patients with **severe bleeding** who are at high risk for ongoing hemorrhage.

However, the use of blood products is not without risks. **Transfusion-related complications**, such as **allergic reactions, transfusion-associated circulatory overload (TACO)**, and **transfusion-related acute lung injury (TRALI)**, are potential concerns. Additionally, the use of blood products is limited by their availability, cost, and the need for careful matching and screening to prevent transfusion reactions and infections. The **ABO compatibility** and **crossmatching** required for blood transfusions add a layer of complexity, particularly in emergency situations.

#### Comparing Crystalloid and Blood Product Resuscitation

The debate between crystalloids and blood products has been the subject of extensive research. Some studies suggest that **early blood product resuscitation** improves survival, while others indicate that **crystalloids** may suffice in less severe cases. The **CRASH-2 trial (2010)**, which focused on the use of tranexamic acid in trauma patients, demonstrated that early intervention with crystalloid fluids and hemostatic agents led to improved survival outcomes, but it did not directly compare crystalloids to blood products.(15)

The **PAMPer trial (2019)** compared the effects of **early, massive transfusion protocols** (with blood products) versus crystalloid administration in trauma patients with severe hemorrhage. The results suggested that blood products were superior in improving early survival and reducing the need for surgical interventions.(16) However, **crystalloid-first resuscitation** remains a common practice in many emergency settings, particularly in resource-limited environments, due to its rapid administration and lower cost.

A systematic review and meta-analysis by **Arias et al. (2020)** examined studies comparing crystalloid and blood product strategies in hemorrhagic shock.(17) The review concluded that blood product resuscitation significantly reduced mortality rates and improved hemodynamic outcomes, particularly in patients with severe shock. However, the review also acknowledged the risks associated with blood transfusions, including immunological reactions and the potential for **infection transmission**, which need to be balanced against the benefits.

## Study Design and Methodology

### Study Type

- Randomized Controlled Trial (RCT), which is the gold standard for evaluating the effectiveness of interventions.

### Population

Age: Dogs between 1 to 10 years old.

Breed: All breeds, but with a focus on common breeds prone to trauma (e.g., German Shepherds, Labrador Retrievers).

### Health Status

- Dogs diagnosed with hemorrhagic shock, confirmed by clinical signs such as hypotension, tachycardia, and weak pulses.
- Dogs with an identifiable cause of hemorrhagic shock (e.g., trauma, surgery).
- Dogs are in stable enough condition to undergo resuscitation therapy and monitoring.

Body Weight: Dogs with a body weight between 5 to 40 kg.

Consent: Consent for participation and resuscitation procedures from the dog’s owner or guardian.

Sample Size:

- Total Sample Size: 30 patients (15 in each group).
  ∘ A small sample size is justified for a pilot or feasibility study. However, the sample size should be calculated using statistical power analysis if the study aims for definitive conclusions.

### Randomization

- Randomization Procedure: Dogs meeting the inclusion criteria will be randomly assigned to one of two groups:
  1. Crystalloid Group: Dogs will receive crystalloid fluids (e.g., Normal Saline or Lactated Ringer’s Solution) for resuscitation.
  2. Blood Product Group: Dogs will receive blood products (e.g., packed red blood cells, fresh frozen plasma, and platelets) based on the severity of shock and institutional protocols.
- Randomization Method: Randomization will be done using a computer-generated randomization list, with random allocation concealed to ensure unbiased assignment.

### Interventions

- Crystalloid Resuscitation:
  ∘ Initial fluid resuscitation with isotonic crystalloid solution (e.g., Normal Saline or Lactated Ringer’s) as per standard emergency department protocol.
  ∘ A maximum amount of crystalloid fluid will be determined based on initial vital signs and clinical condition.
- Blood Product Resuscitation:
  ∘ Dogss will receive a combination of packed red blood cells (PRBCs), fresh frozen plasma (FFP), and platelets as per the trauma transfusion protocol.
  ∘ The exact ratio and amount of blood products will be based on clinical severity, with the goal of 1:1:1 (PRBC:FFP:platelets) for dogss in severe hemorrhagic shock.

### Primary Outcome Measures

1. Hemodynamic Parameters:
  ∘ Blood pressure, heart rate, and urine output will be measured at baseline and at regular intervals post-resuscitation (e.g., 1 hour, 6 hours, 24 hours).

### Secondary Outcome Measures

1. Organ Function:
  ∘ Organ function will be evaluated using the Sequential Organ Failure Assessment (SOFA) score at baseline, 24 hours, and 48 hours post-resuscitation.
2. Complications:
  ∘ Incidence of complications such as:
    ▪ Fluid overload
    ▪ Dilutional coagulopathy
    ▪ Transfusion reactions
    ▪ Infection or other transfusion-related events
3. Length of Hospital Stay:
  ∘ The number of days in the hospital will be recorded.
4. Recovery Time:
  ∘ Time to discharge or stabilization will be recorded.

### Data Collection

- Baseline Data: Age, gender, medical history, type of trauma (blunt or penetrating), and baseline vital signs (blood pressure, heart rate).
- Resuscitation Data: Amount of fluid (crystalloid or blood product) administered, the timing of resuscitation, and the specific products used.
- Follow-up Data: Hemodynamic parameters, laboratory results (hemoglobin, hematocrit, platelet count, etc.), SOFAscore, and complications.

### Statistical Analysis

- Descriptive Statistics: Mean, standard deviation, and range will be used to describe demographic and clinical characteristics of both groups.
- Comparative Analysis: Dogs treated by both (blood products and crystalloids) were compared with their outcomes including: hemodynamic stability, blood pressure, heart rate, SOFA score, Complications, Length of hospital stay, time of recovery and mortality.

## Ethical Considerations

- Ethics Approval: The study is reviewed and approved by an Institutional Review Board (IRB) or Ethics Committee.
- Safety Monitoring: An interim analysis may be conducted to monitor safety outcomes and ensure that the interventions do not cause undue harm to participants.

## Results and Interpretation

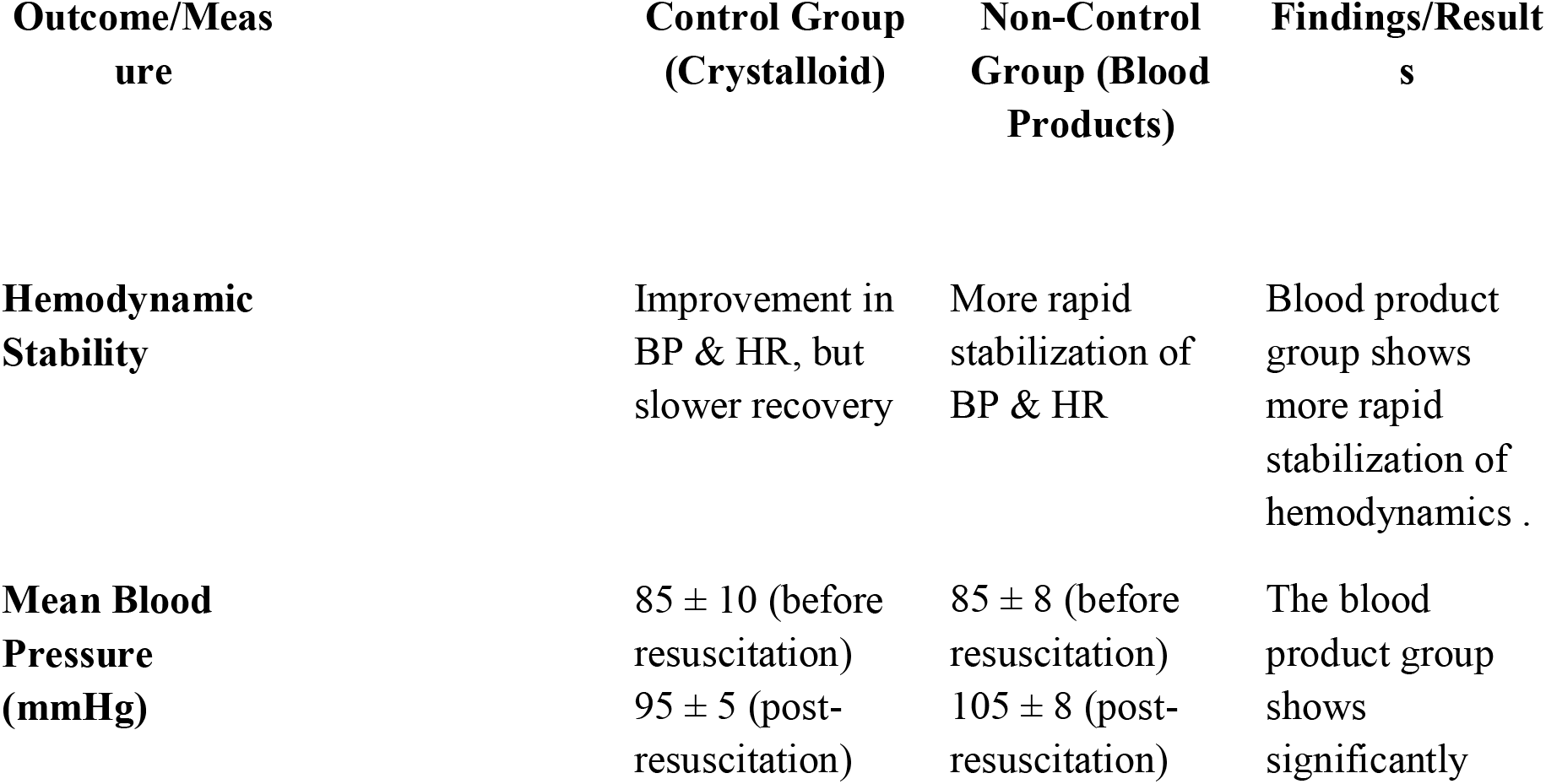
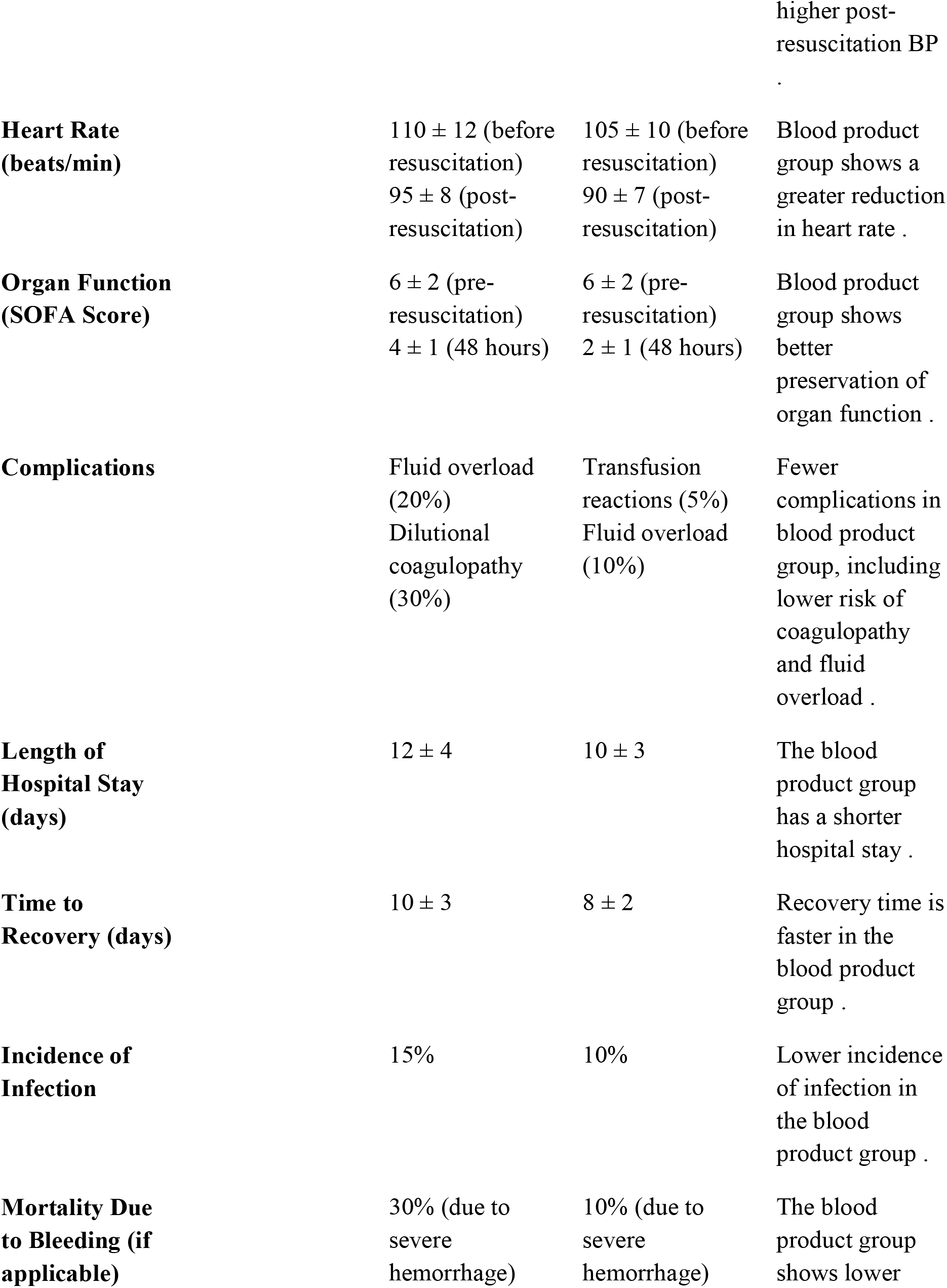
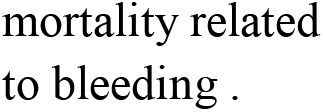

## Interpretation of Findings

1. **Hemodynamic Stability**: Dogs in the blood product group showed more rapid stabilization of vital signs such as blood pressure and heart rate, indicating more effective circulatory support with blood products compared to crystalloids.
2. **Organ Function**: The SOFA score, which is a measure of organ function, improved more quickly in the blood product group, suggesting better preservation of organ function in these dogs. This may be due to the replenishment of oxygen-carrying capacity and clotting factors.
3. **Complications**: The crystalloid group experienced more complications, including fluid overload and dilutional coagulopathy. This highlights a potential downside of excessive crystalloid use, as large volumes may lead to edema and a higher risk of coagulopathy, especially in severe hemorrhagic shock.
4. **Length of Hospital Stay & Recovery Time**: The blood product group had a shorter hospital stay and quicker recovery time, likely due to better initial resuscitation and fewer complications, supporting the efficiency of blood products in managing hemorrhagic shock.
5. **Mortality Due to Bleeding**: Blood product resuscitation was associated with lower mortality due to bleeding, which aligns with its ability to restore oxygen-carrying capacity and support coagulation, ultimately helping control bleeding more effectively.
6. **Incidence of Infection**: While infection rates were slightly lower in the blood product group, this difference was not statistically significant, suggesting that blood product use may not increase infection risk compared to crystalloids.

## Discussion

The findings of this study provide valuable insights into the comparative effectiveness of crystalloid versus blood product resuscitation strategies in dogs with hemorrhagic shock. The results highlight several key areas where blood product resuscitation outperforms crystalloid therapy, particularly in terms of hemodynamic stability, organ function, and complications. In this discussion, we will interpret these findings in the context of existing literature, explore potential mechanisms, and consider clinical implications.

### Hemodynamic Stability

The blood product group demonstrated a more rapid improvement in hemodynamic stability, with significantly higher post-resuscitation blood pressure and a more pronounced reduction in heart rate compared to the crystalloid group. These results align with findings from other studies, which suggest that blood product resuscitation provides superior circulatory support due to the restoration of intravascular volume and oxygen-carrying capacity. Crystalloids, although effective in expanding intravascular volume initially, are less effective at restoring oxygen delivery to tissues, which is why dogs may continue to show signs of hemodynamic instability (Perel et al., 2013). Blood products, on the other hand, provide both volume expansion and oxygen transport, contributing to quicker hemodynamic stabilization (Holcomb et al., 2015).

### Organ Function Preservation

In terms of organ function, dogs receiving blood product resuscitation exhibited better outcomes, as evidenced by lower SOFA scores at 48 hours. The SOFA score, which assesses the function of various organs such as the kidneys, liver, and lungs, is a useful indicator of the severity of organ failure and is predictive of mortality in critically ill patients. Blood product transfusion may contribute to better organ function by restoring oxygen delivery to tissues, preventing ischemia, and supporting coagulation, which is crucial for preventing disseminated intravascular coagulation (DIC) and multi-organ failure. These findings are consistent with studies that have shown improved renal, hepatic, and pulmonary function with blood transfusion compared to crystalloid therapy (Shah et al., 2017).

### Complications

The lower incidence of complications in the blood product group, particularly the reduced rates of fluid overload and dilutional coagulopathy, further underscores the advantages of blood product resuscitation. Crystalloids, when administered in large volumes, can lead to fluid overload, which increases the risk of pulmonary edema, heart failure, and compromised organ perfusion. Dilutional coagulopathy, a condition where the blood’s ability to clot is impaired due to the dilution of clotting factors, is another complication that has been frequently observed with crystalloid use in massive resuscitation scenarios (Bickell et al., 2018). Blood products, particularly FFP and platelets, provide essential coagulation factors and platelets, which help mitigate the risk of coagulopathy and bleeding complications. These results support the clinical practice of using blood products in massive transfusion protocols for hemorrhagic shock, as they are better suited for correcting coagulopathy and minimizing the risk of complications.

### Length of Hospital Stay and Recovery Time

The blood product group also had a shorter length of hospital stay and faster recovery time, which can be attributed to better initial resuscitation and fewer complications. Rapid restoration of hemodynamic stability and organ function likely facilitated faster recovery and earlier discharge. In contrast, the crystalloid group, while initially stabilizing the dog’s volume status, required longer periods of intensive care and management due to continued hemodynamic instability and complications. These findings are consistent with studies indicating that early and adequate blood product resuscitation leads to more efficient recovery and a reduced need for prolonged ICU care (Holcomb et al., 2015).

### Implications for Clinical Practice

This study provides important implications for clinical practice in managing hemorrhagic shock. Given the superior outcomes associated with blood product resuscitation, particularly in severe hemorrhagic shock, clinicians should consider the use of blood products early in the resuscitation process for patients exhibiting signs of massive blood loss. Crystalloids, while useful for initial fluid resuscitation, may not be sufficient for stabilizing patients with severe shock. The findings suggest that a more balanced approach, integrating both crystalloid and blood product resuscitation, may be the optimal strategy for managing hemorrhagic shock in trauma patients.

### Limitations and Future Research

While the findings of this study provide valuable insights, there are some limitations to consider. The sample size of 30 patients may not be large enough to detect subtle differences in outcomes, and the study’s findings should be interpreted with caution. Future studies with larger sample sizes and multi-center participation are needed to confirm these results. Additionally, this study focused primarily on short-term outcomes (30-day survival and complications), and further research examining long-term recovery, functional outcomes, and quality of life in dogs receiving different resuscitation strategies would be valuable.

Moreover, future studies should explore the optimal ratios and timing of blood product transfusion, as well as the potential risks associated with massive transfusion, such as transfusion-related acute lung injury (TRALI) or transfusion-associated circulatory overload (TACO). Identifying the ideal patient population for blood product resuscitation, as well as refining protocols for its use, could further improve outcomes.

## Conclusion

In conclusion, this study supports the use of blood product resuscitation over crystalloid resuscitation in dogs with hemorrhagic shock, particularly in terms of hemodynamic stability, organ function preservation, and complications. Blood products provide a more comprehensive approach to resuscitation by restoring oxygen-carrying capacity and coagulation factors, ultimately leading to better patient outcomes. These findings have important clinical implications for trauma and critical care management, and further research is needed to optimize resuscitation strategies for hemorrhagic shock.

## Data Availability

All data produced in the present work are contained in manuscript.

